# One Day Hospital Initiation of Oral Sotalol The Cmax ss Test Strategy

**DOI:** 10.64898/2026.03.12.26348293

**Authors:** Janos Molnar, John C. Somberg

**Affiliations:** American Institute of Therapeutics, 21 N Skokie Hwy, Suite G-3, Lake Bluff, IL 60044, USA

**Keywords:** Sotalol, loading, proarrhythmia, QT prolongation

## Abstract

**BACKGROUND:** Sotalol loading intravenously enables achieving blood levels of sotalol that are observed at maximal steady-state concentration (Cmax ss) in one-day permitting the measurement of maximum QTc effects. Rapid evaluation of the QTc effects permits determination of arrhythmic risk and thus permits discharge in 24-hours instead of the usual three-day oral load hospitalization. Given the expense of IV Sotalol an oral loading test strategy is presented that also achieves Cmax ss blood levels rapidly, permitting a one-day hospitalization for QTc evaluation.

**METHOD:** Pharmacokinetic parameters referred to in the literature derived from normals as well as patients was utilized for population pharmacokinetic modeling and simulation.to obtain the Cmax ss concentrations for patients with normal renal function, creatinine clearance (CrCl) > 90 ml/min), as well as for patients with a CrCl of 60-89, 30-59, and 10-29 ml/min). Using pharmacokinetic simulations, an oral loading dose, as well as a second oral dose were determined that would reach the estimated Cmax ss in each of the groups based on renal function.

**RESULTS:** For target dosing of 120 mg oral sotalol BID in patients with a CrCl >90 ml/min an oral loading dose of 200 mg provides a peak sotalol level of 1,420 ng/ml in 3-4 hours post dosing. The Cmax ss target is 1,299 ng/ml resulting in a 9% overshoot. The Cmax ss concentration provides a means of evaluating QTc effects within 24-hours. Oral loading regimens are described for varying additional renal function levels (CrCl 60-90, 30-59 and 10-29 ml/min) along with the time to first oral dose and follow-up dosing. The initial test dose can be based on an 80 or 120 mg oral sotalol maintenance dosing strategy.

**CONCLUSIONS:** Employing an oral loading strategy may permit QTc evaluation and one-day discharge, preserving the pharmacoeconomic advantage of a Cmax ss test strategy.

**Clinical Perspective:** *What is Known?:*

- Intravenously loading of sotalol enables achieving blood levels that are observed at maximal steady-state concentration (Cmax ss) in one-day permitting the measurement of maximum QTc effects.
- Rapid evaluation of the QTc effects permits determination of arrhythmic risk and thus permits discharge in 24-hours instead of the usual three-day oral load hospitalization

*What the Study Adds:*

- With oral sotalol loading, the Cmax ss can also be achieved in one-day permitting the measurement of maximum QTc effects and discharge from the hospital in 24-hours instead of the usual three-day inpatient initiation of oral sotalol.

## Introduction

Sotalol is a Type III anti-arrhythmic pharmaceutical that prolongs the action potential duration (APD) by prolonging repolarization of cardiac cells, [1,2] as well as a beta blocker. The prolongation of APD is the primary mechanism of anti-arrhythmic action as well as the underlying cause of facilitation of the life-threatening ventricular arrhythmias, termed pro-arrhythmia. Pro-arrhythmia may occur at a rate of 3 to 6% of patients taking sotalol therapy, but when it occurs may lead to a fatal arrhythmias.[2,3,4,5] Sotalol is frequently employed to prevent the recurrence of atrial fibrillation.[6] Recent guideline revisions have downgraded oral sotalol use from a first line anti-arrhythmic to a second line choice. (refs) The potential for pro-arrhythmia requires that sotalol be evaluated for excessive QT prolongation. The FDA recommends in the product insert that the evaluation of pro-arrhythmia be done through a three-day hospitalization where oral sotalol is slowly loaded till the blood concentration of sotalol reaches its maximal steady state concentration (Cmax ss). [2] By undertaking the oral loading procedure one can assess the extent of QT prolongation and thus can avoid administering sotalol to those patients showing an excessive degree of QT prolongation. Recently Somberg described an IV loading procedure that shortens the in-hospital loading by utilizing an IV formulation of sotalol, reducing the administration time from 3 days to 1 day of in-hospital observation.[7] This loading regimen has been confirmed in patient studies in the PEAKS Registry. (all 6 refs) The shortening of the observation period is most convenient for patients, as well as possibly saving 2 days of hospitalization costs and permitting hospitals to utilized telemetry beds for other purposes, a benefit since telemetry beds in-hospital are usually in scarce availability.

Others have advocated a combined oral and IV administration of sotalol to reach a steady-state maximum concentration in patients undergoing in-hospital monitoring.[8,9,10,11] With the ever-increasing cost of IV sotalol, the cost approaching $4,000 per unit, the pharmacoeconomic benefit from cost savings is gone, causing physicians and hospitals to return to the three-day loading which is most inconvenient for patients, or initiating sotalol on an outpatient basis that entails the risk of QT prolongation and proarrhythmia unsupervised.

To permit a one-day in-hospital loading regimen to determine safety of long-term administration of sotalol, an oral loading regimen for sotalol is proposed that can be adjusted for different levels of renal function.

## Methods

Pharmacokinetic parameters referred to in the literature derived from normals as well as patients was utilized for population pharmacokinetic (PK) modeling and simulation.to obtain the Cmax ss concentrations.[7] A detailed publication of the modeling methodologies are reported elsewhere.[12] The Cmax ss was calculated for patients with normal renal function (greater than 90 ml/min), as well as for patients with a CrCl of 60-90, 30-59, and 10-29 ml/min). Using the WinNonlin, Version 5 program an oral loading dose was determined that would reach the estimated Cmax ss in approximately 3-4 hours. Then a second oral dose was calculated that when administered at a selected interval from the first oral dose will reach Cmax ss a second time. Following this the time to next dose as well as the dose selected for maintenance therapy was determined that would not exceed Cmax ss. Using this approach the effect of oral dosing could be determined on the QTc electrocardiograph interval within 24-hours to determine if there will be excessive QTc prolongation or the effects on the QTc are within an acceptable range as specified in the sotalol package insert (less than an increase of 500 msec). Simulations showing time course of sotalol concentrations following a 1^st^ oral loading dose, 2^nd^ oral dose and follow up maintenance dosing was constructed for each of the designated renal function groups.

## Results

Sotalol is usually administered twice daily based on a half-life of 12-hours. The maintenance dose of sotalol is adjusted for renal function since elimination is through renal clearance. The dosing concentration is determined by the effect on the QTc interval duration with the product insert carrying a black box warning with the direction that sotalol not be initiated in patients with a QTc longer than 450 msec. Additionally, the prescriber is warned that if the QTc interval prolongs to 500 msec or greater on sotalol the dose of sotalol must be reduced or its administration stopped.

### Target Maintenance 120 mg (CrCl 90 ml/min or greater)

The most frequently employed dose of sotalol is 120 mg PO every 12 hours. For a normal creatinine clearance (CrCl of 90 ml/min or greater) the Cmax ss has been found to be on average 1,299 ng/ml. Simulations have shown that a 200 mg oral dose of sotalol will on average result in a serum concentration of 1,420 ng/ml resulting in a 9% overshoot. If the plan is to use a maintenance dosing schedule with 120 mg oral sotalol dose, then 120 mg of sotalol can be administered 12 hours after the first oral loading dose. During the administration of the 1^st^ and 2^nd^ sotalol doses the patient’s QTc can be monitored to avoid excessive QTc prolongation. If the QTc is not excessively prolonged then the patient can be discharged from ECG monitoring in hospital and oral sotalol maintenance dosing 120 mg every 12 hours thereafter. The dosing simulation can be seen in Figure 1.

**Figure 1.**
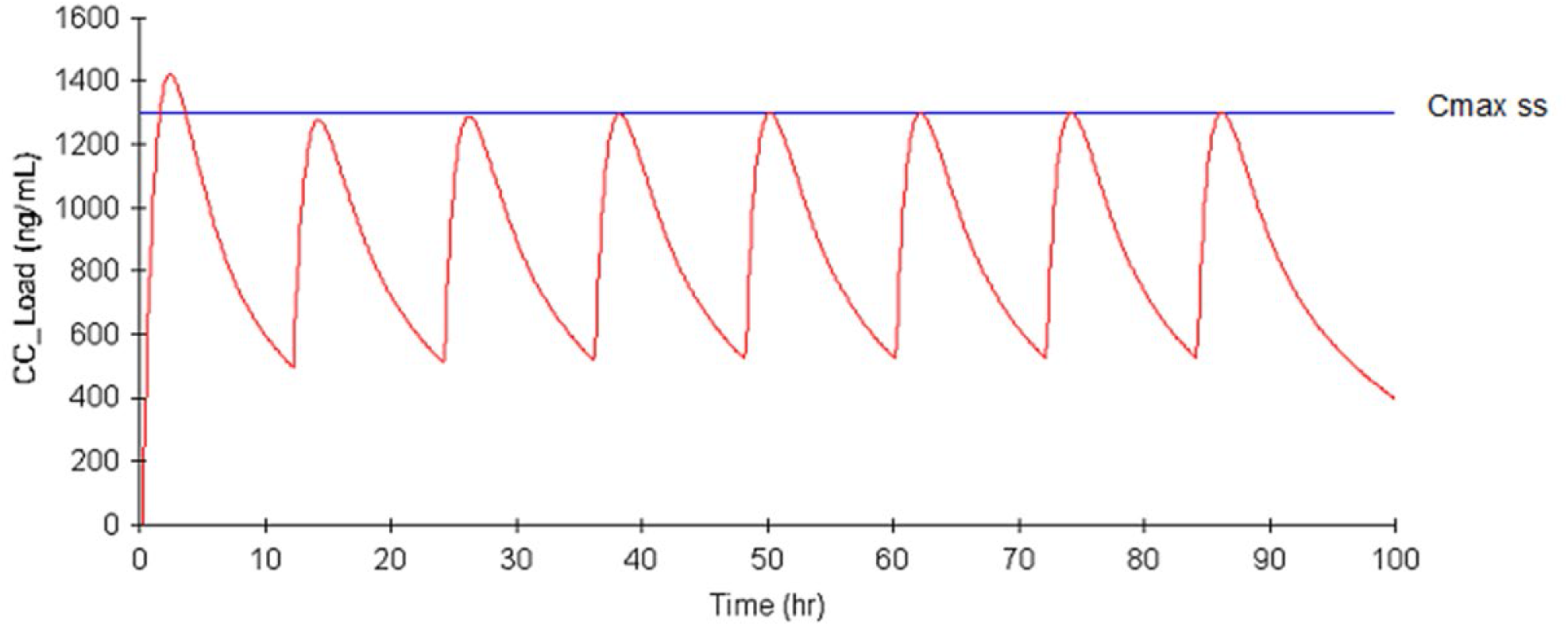
Loading for target dosing of 120 mg oral sotalol BID in patients with normal renal function (Creatinine clearance >90 ml/min). The first oral dose is 200 mg followed 12 hours by 120 mg oral sotalol and then every 12 hours thereafter. The read line indicates the mean sotalol plasma concentration and after chronic dosing. The Cmax is 1420 ng/ml and the Cmax ss is 1299 ng/ml with a 9% loading overshoot. The blue line indicates the mean maximal steady state (Cmax ss) plasma concentration.

### Target Maintenance 80 mg Dose (CrCl of 90 ml/min or greater)

The steady-state maximum concentration administered with oral sotalol dosing of 80 mg twice daily results in a concentration of 862 ng/ml. For an 80 mg target maintenance dosing the loading dose would be 120 mg followed by 80 mg bid (see Fig. 2).

**Figure 2.**
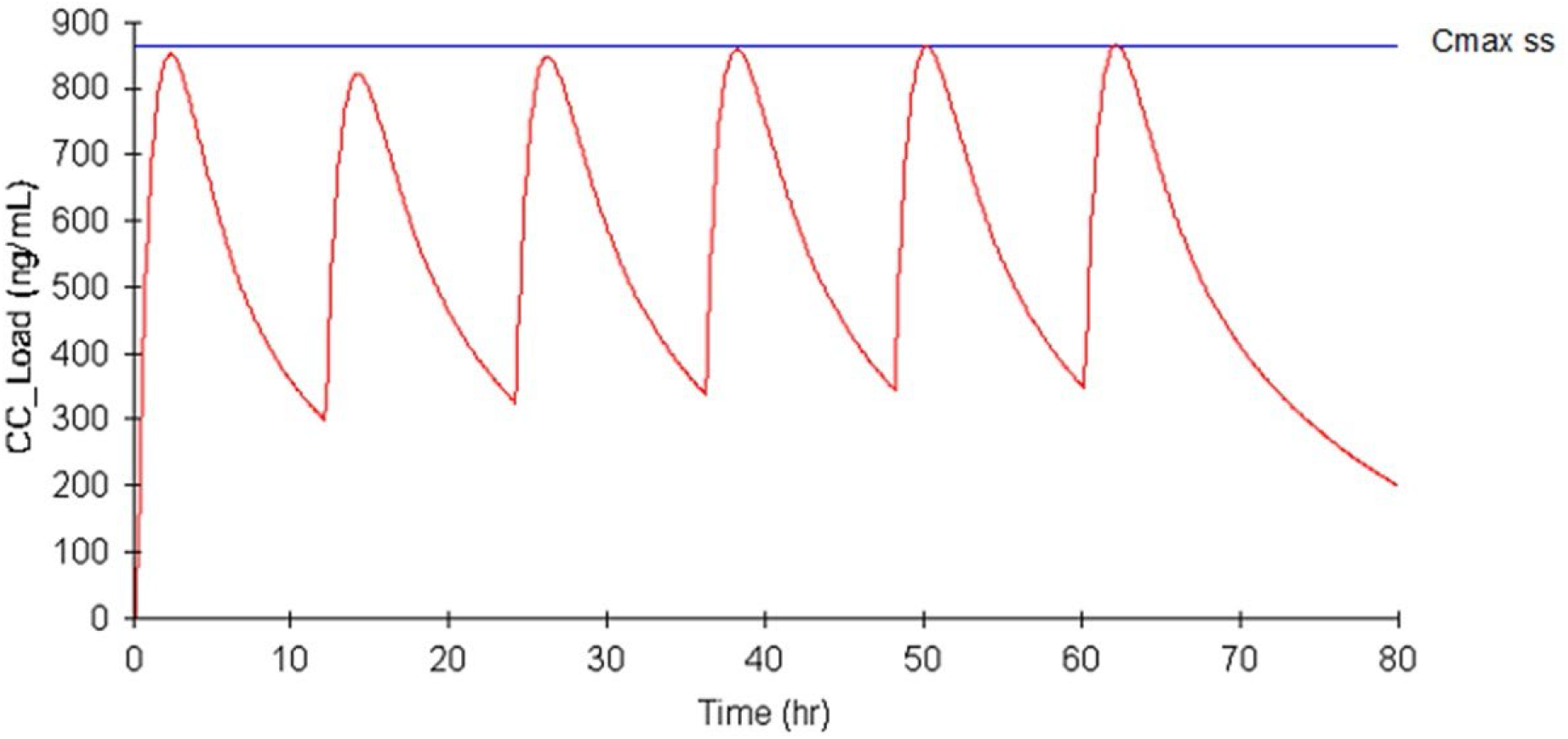
Loading for target dosing of 80 mg oral sotalol BID in patients with normal renal function (Creatinine clearance >90 ml/min). The first oral dose is 120 mg followed by 80 mg oral sotalol 12 hours after the first dose and then taken every 12 hours thereafter. The loading dose Cmax is 851 ng/ml and the Cmax ss is 862 ng/ml.

### Target Maintenance Dose 120 mg CrCl 60-90 ml/min

With a CrCl between 60-90 ml/min and the oral loading dose would be 240 mg followed by 120 mg 8 hours after the first oral loading dose and the maintenance dosing of 120 mg every 12 hours thereafter. The resulting Cmax would be 1,889 ng/ml following the first oral dose. With the target Cmax ss of 1,800 ng/ml with chronic oral dosing the resulting overshoot would be 5%. (Fig. 3)

**Figure 3.**
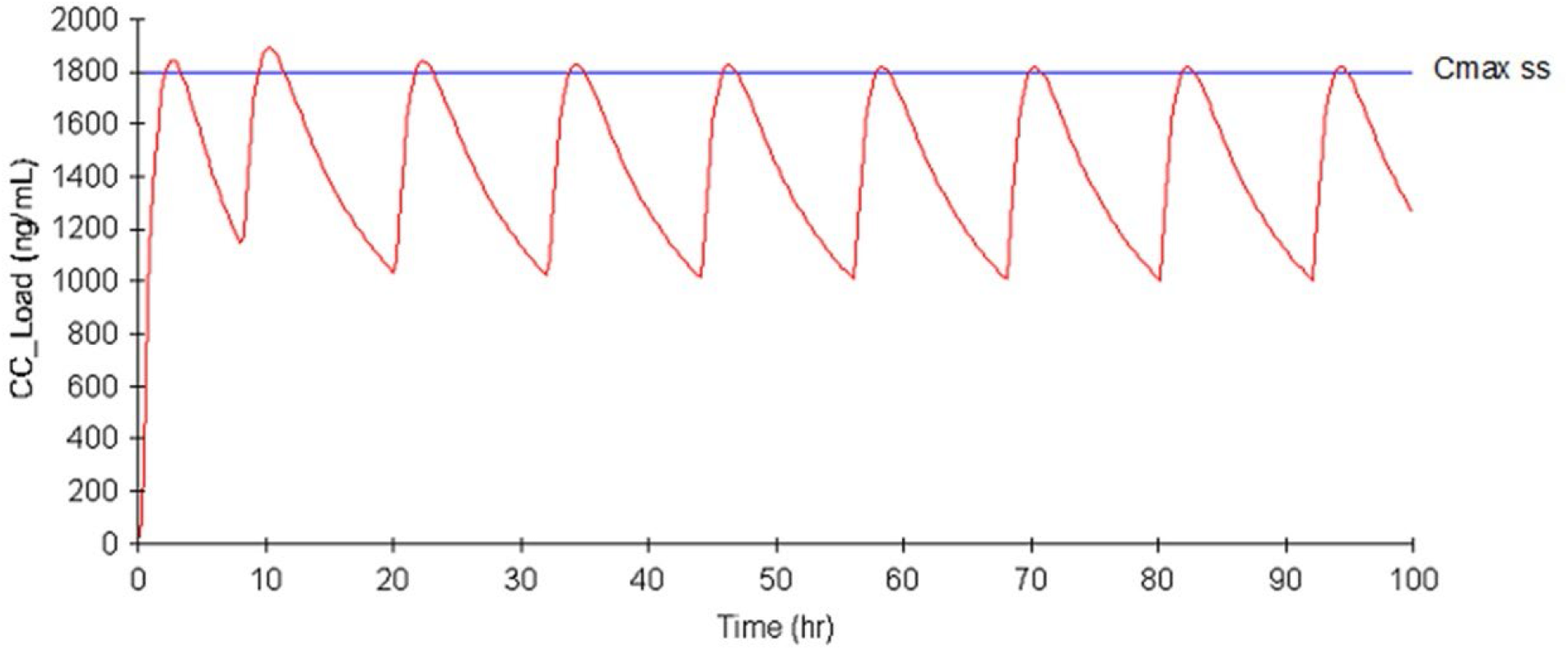
Loading for target dose of 120 mg oral sotalol BID in patients with Creatinine clearance between 60 and 90 ml/min. The loading oral dose is 240 mg followed by 120 mg oral 8 hours after the first dose and then 120 sotalol taken every 12 hours. The Cmax is 1889 ng/ml and the Cmax ss is 1800 ng/ml resulting in an initial overshoot of 5%.

### Targeting a Maintenance Dose of 80 mg CrCl 60-90 ml/min

With a CrCl between 60-90 ml/min and the oral maintenance target of 80 mg twice daily, the oral loading dose would be 160 mg yielding a concentration of 1,220 ng/ml. The second dose can be followed after 8 hours by 80 mg orally and then 80 mg bid thereafter. The concentration reached by the first oral dose would be 1,271 ng/ml. Since the Cmax ss is 1,220 ng/ml with chronic oral dosing the loading dose will result in a 4% overshoot. (see Fig 4).

**Figure 4.**
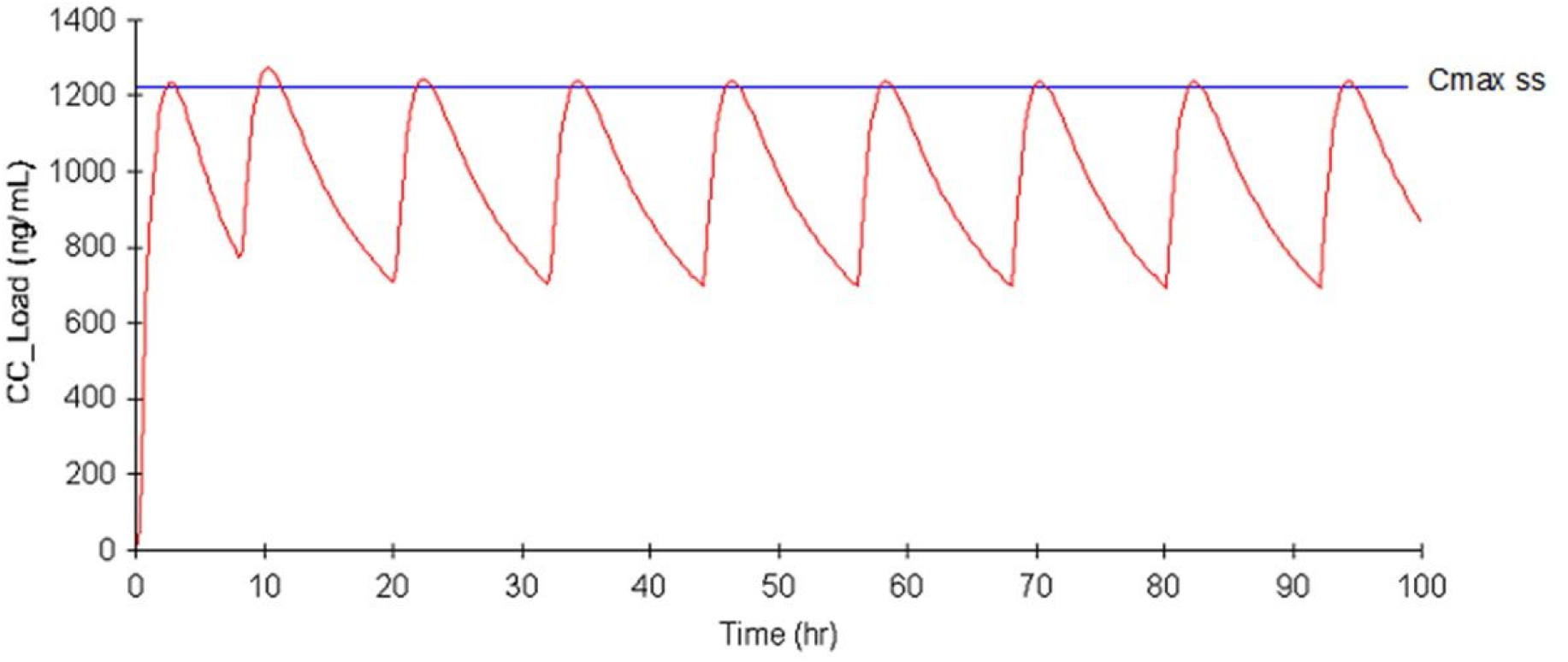
Loading for target dose of 80 mg oral sotalol BID in patients with Creatinine clearance between 60 and 90 ml/min. The loading dose is 160 mg followed by 80 mg oral sotalol at 8 hours after the first dose and then 80 sotalol every 12 hours. The Cmax is 1271 ng/ml and the Cmax ss is 1220 ng/ml resulting in an initial 4% overshoot.

### Targeting a Maintenance Dose of 120 mg, CrCl 30-59 ml/min

Patients with a creatinine clearance of between 30-59 ml/min with a target maintenance dose 120 mg, the loading dose would be 220 mg orally yielding a Cmax of 1,785 ng/ml. Since the average Cmax ss is 1,620 ng/ml with chronic oral dosing the estimated overshoot would be 10%. The second oral dose should be administered 24 hours later followed by an oral dose of 120 mg and 120 mg every 12 hours thereafter. (see Fig 5)

**Figure 5.**
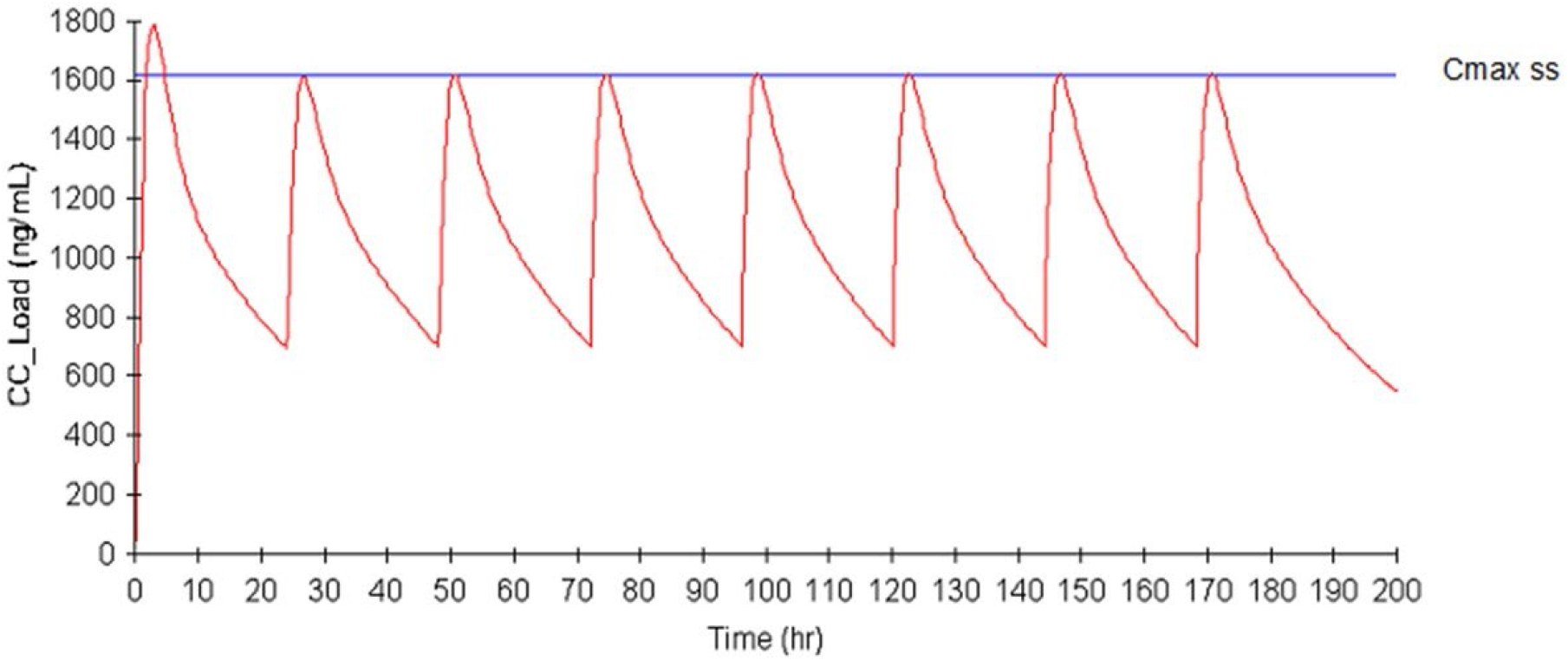
Loading for a target maintenance dose of 120 mg BID in patients with Creatinine clearance between 30 and 59 ml/min. The loading dose is 220 mg followed by 120 mg sotalol taken every 24 hours. The Cmax after the loading dose is 1785 ng/ml and Cmax ss is 1620 resulting in a 10% overshoot.

### Targeting a Maintenance Dose of 80 mg, CrCl 30-59 ml/min

For patients with a CrCl 30-59 ml/min selected to receive a maintenance dose of 80 mg, the initial loading dose would be 140 mg that would yield a Cmax of 1,138 ng/ml on average. Since the estimated Cmax ss is 1,090 ng/ml with chronic oral dosing one would see an overshoot of 4%. The second oral dose of 80 mg can be administered in 24 hours and then 24 hours thereafter. (see Fig. 6)

**Figure 6.**
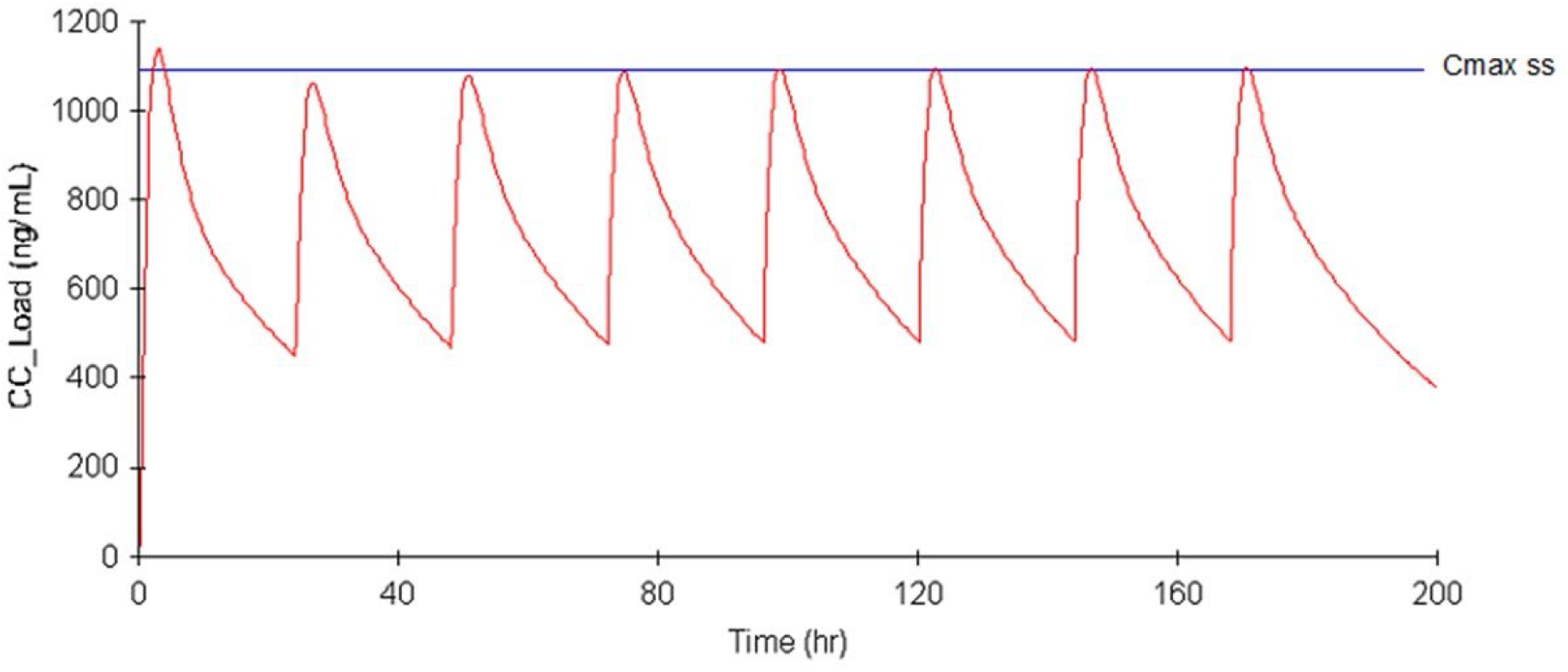
Loading for a target dose of 80 mg BID in patients with Creatinine clearance between 30 and 59 ml/min. The first oral dose is 140 mg followed by 80 mg sotalol taken every 24 hours. The Cmax after first dose is 1138 ng/ml and Cmax ss is 1090 resulting in a 4% overshoot.

### Targeting Maintenance Dosing of 120 mg CrCl 10-29 ml/min

Patients with a renal function of 10-30 ml/min with a targeted maintenance dose of 120 mg could receive a loading dose of 200 mg that would result in a Cmax of 1,706 ng/ml. With the Cmax ss estimate of 1,610 ng/ml with chronic oral dosing an overshoot of 6% would occur. The second oral dose could be administered in 48 hours and then 120 mg every 48 hours thereafter. (see Fig 7)

**Figure 7.**
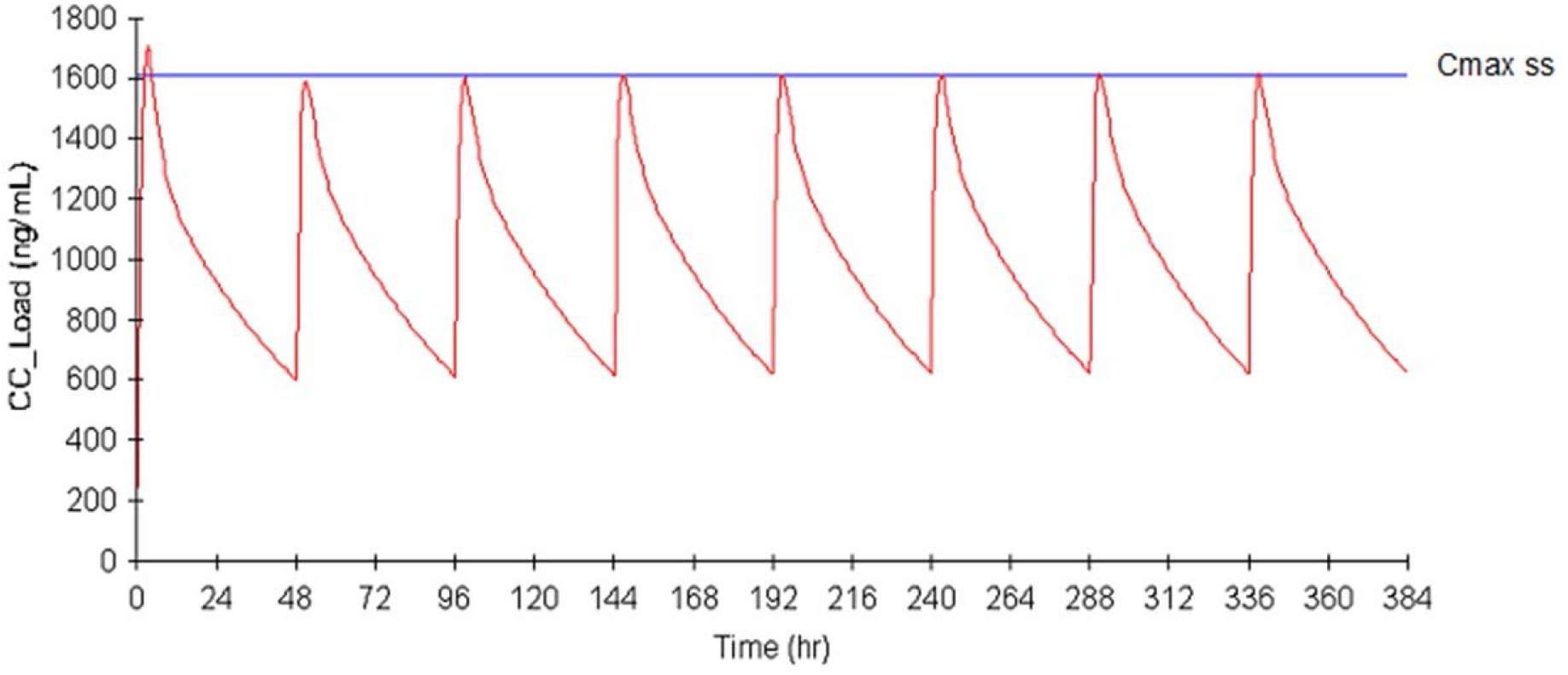
Loading for a target of 120 mg BID in patients with Creatinine clearance between 10 and 29 ml/min. The first oral dose is 200 mg followed by 120 mg sotalol taken every 48 hours. The Cmax after first dose is 1706 ng/ml and the Cmax ss is 1610 ng/ml, a 6% overshoot.

### Targeting Maintenance Dosing of 80 mg CrCl 10-29 ml/min

Patients with a renal function of 10-30 ml/min CrCl with a targeted maintenance dose of 80 mg the loading dose of 140 mg would yield a Cmax of 1,196 ng/ml. With a Cmax ss of 1,090 ng/ml with chronic oral dosing a 10% overshoot would be seen. The second oral dose of 80 mg would be administered 48 hours later and then 80 mg every 48 hours to follow. (see Fig 8)

**Figure 8.**
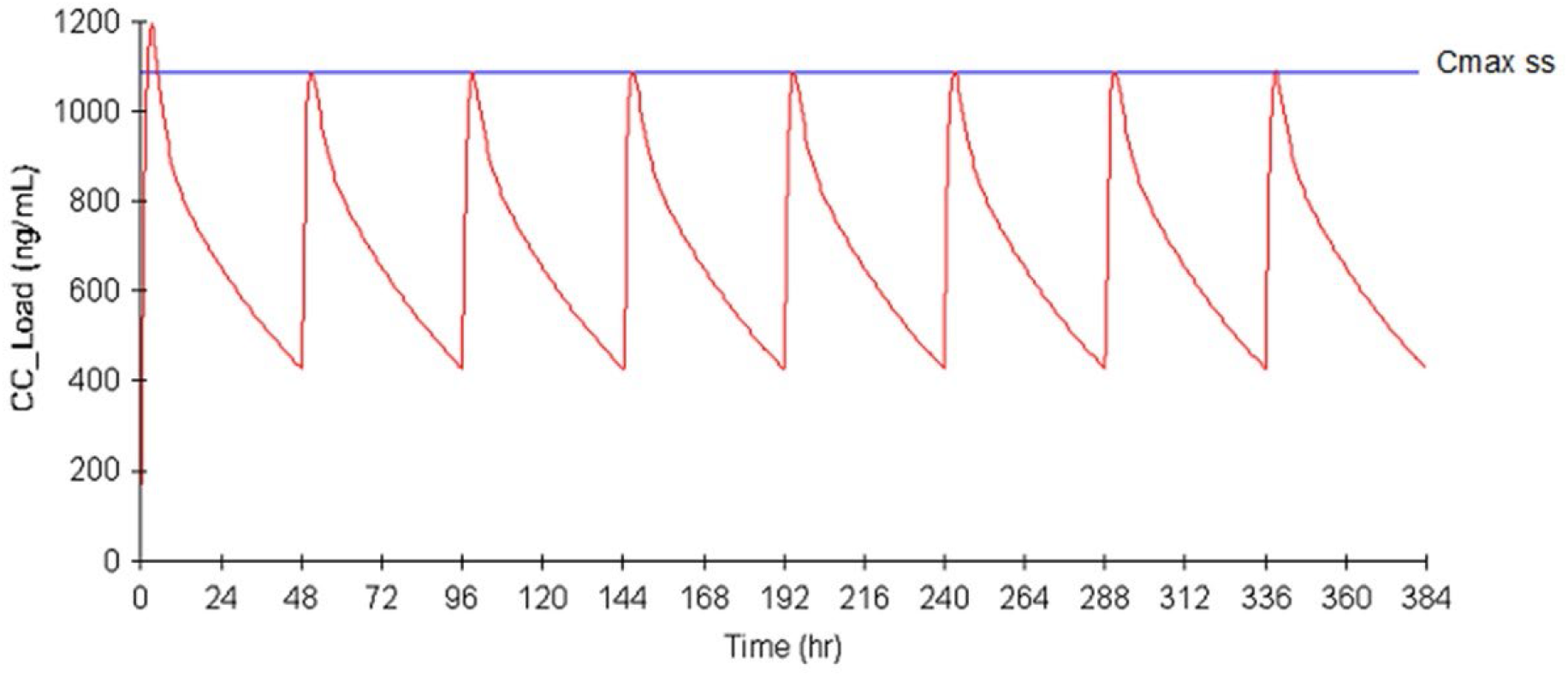
Loading for a target dose of 80 mg oral sotalol BID in patients with Creatinine clearance between 10 and 29 ml/min. The first oral dose is 140 mg followed by 80 mg sotalol taken every 48 hours. The Cmax after the first dose is 1196 ng/ml and the Cmax ss is 1090 ng/ml resulting in a 10% overshoot.

## Discussion

Physicians who decide to utilize oral sotalol, a second line anti-arrhythmic therapy need to select a target maintenance dose that they believe will be an effective anti-arrhythmic dosing regimen. The physician next needs to determine the patient’s renal function to determine the maintenance dosing interval. Then the effect of sotalol on QTc interval prolongation needs to be determined. Since some patients despite having the appropriate dose calculated based on renal function can still have excessive QTc prolongation that could result in serious pro-arrhythmia, the effect of sotalol exposure on the QTc interval needs to be ascertained by ECG monitoring in-hospital. For patients with normal renal function or those with slightly decreased function a 3-day in-hospital load can assess the QTc effect. Employing a loading strategy can attenuate the observation period from 3-days to 1-day utilizing an IV loading strategy. Using IV loading has the advantage of being able to stop the infusion if the QTc prolongation becomes excessive or if arrhythmias develop. However, the high cost of IV Sotalol makes this approach economically difficult.

Oral sotalol loading can also achieve an initial sotalol blood concentration that reaches a peak concentration that is seen with chronic oral dosing with a one-day hospital observation. The advantage is the low cost of oral sotalol, but the disadvantage is that once the oral dose is administered the sotalol administration can’t be stopped if the QTc prolongs excessively and/or arrhythmias develop. In the patients with increased risk factors for excessive QTc prolongation the oral loading approach may not be appropriate, but for most patients this approach seems reasonable though it has not been reviewed or approved by the FDA.

In patients with normal or slightly decreased renal function the first and second oral dose effects on the QTc can be observed in 24-hours of hospitalization. Those patients with renal function of 30-59 and 10-29 ml/min can have the effect of the 1^st^ oral dose on the QTc assessment in 24-hours, but the 2^nd^ dose effect would occur after 24-hours. Depending upon patient risk, as evaluated by the prescribing physician, the second dose may need to prolong hospitalization beyond 24-hours. But this is also true for IV Sotalol. In patients with reduced renal function and other factors such as a prior MI, low EF or known propensity to bradycardia, in-hospital loading IV with the ability to stop the infusion if the QTc prolongs excessively may be a safer approach. We are not recommending choosing sotalol for chronic therapy in a given patient, but rather providing an alternative oral loading approach that may be cost effective and convenient for the patient.

### Limitations

The sotalol concentrations presented are mean value and there will be individual variations with outliers that could be due to drug interactions, absorption variability, or food effects. The effect of a given concentration on the QT interval and underlying cardiac repolarization could change with ischemia, myocardial damage, electrolyte changes, deteriorating heart function, or bradycardia increasing the risk of pro-arrhythmia. Predicting long term safety of sotalol on the basis of a one-time evaluation of what is believed to be the highest sotalol concentration (Cmax ss) may not be reliable given the many factors involved. The initial evaluation is just a start and continued monitoring of the patient is required to maintain avoidance of pro-arrhythmias from the drug.

The proposed oral loading regimen is based on pharmacometrics calculations and is not based on clinical studies. However, the dosing recommendation for IV loading are also based on pharmacometric calculations that were approved by the FDA and only recently corroborated by clinical studies. (refs) The same methodology for the oral loading dose selection was employed as was employed for the IV loading methodology and this approach was approved by the FDA for IV sotalol and is incorporated in the product insert.

The Cmax ss dose effect on QTc is not the only factor that may relate to the possibility of pro-arrhythmia. Bradycardia can facilitate Torsade and thus continued observation over the chronic course of therapy utilizing outpatient monitoring ECG techniques is needed to evaluate the potential for adversity.

## Conclusion

Oral sotalol can be utilized as a loading dose to evaluate QTc effects in-hospital with a 24-hour initial hospitalization for safety assessment. Loading with oral sotalol in many patients may be an alternative approach to IV Sotalol loading and preferable to starting sotalol therapy on an outpatient basis.

## Data Availability

Data are not available

## Acknowledgement

none

## References

[1] Singh BN, Vaughan Williams EM. A third class of anti-arrhythmic action. Effects on atrial and ventricular intracellular potentials, and other pharmacological actions on cardiac muscle, of MJ 1999 and AH 3474. Br J Pharmacol. 1970 Aug;39(4):675–87.

[2] Sotalol hydrochloride tablet prescribing information. Available at: https://dailymed.nlm.nih.gov/dailymed/drugInfo.cfm?setid=62a71c8c-d92d-4122-848f-9ae35f8bbe2b.

[3] MacNeil DJ, Davies RO, Deitchman D. Clinical safety profile of sotalol in the treatment of arrhythmias. Am J Cardiol. 1993;72(4):44A–50A. doi: 10.1016/0002-9149(93)90024-7.

[4] Kühlkamp V, Mermi J, Mewis C, Seipel L. Efficacy and proarrhythmia with the use of d,l-sotalol for sustained ventricular tachyarrhythmias. J Cardiovasc Pharmacol. 1997;29(3):373–81. doi: 10.1097/00005344-199703000-00011.

[5] Soyka LF, Wirtz C, Spangenberg RB. Clinical safety profile of sotalol in patients with arrhythmias. Am J Cardiol. 1990 Jan 2;65(2):74A-81A; discussion 82A-83A. doi: 10.1016/0002-9149(90)90207-h.

[6] January CT, Wann LS, Alpert JS, Calkins H, Cigarroa JE, Cleveland JC Jr, Conti JB. et al. 2014 AHA/ACC/HRS guideline for the management of patients with atrial fibrillation: a report of the American College of Cardiology/American Heart Association Task Force on practice guidelines and the Heart Rhythm Society. Circulation. 2014;130(23):e199–267. doi: 10.1161/CIR.0000000000000041.

[7] Somberg JC, Vinks AA, Dong M, Molnar J. Model-Informed Development of Sotalol Loading and Dose Escalation Employing an Intravenous Infusion. Cardiol Res. 2020;11(5):294–304. doi: 10.14740/cr1143.

[8] Varela DL, Burnham TS T, May HL Bair T, Steinberg BAB Muhlestein JL Anderson JU Knowlton K, Jared Bunch T. Economics and outcomes of sotalol inpatient dosing approaches in patients with atrial fibrillation. J Cardiovasc Electrophysiol. 2022;33(3):333–342. doi: 10.1111/jce.15342

[9] Liu AY, Charron J, Fugaro D, Spoolstra S, Kaplan R, Lohrmann G, Gao X, Gay H, Passman R, Kim S, Lin AC, Chicos A, Arora R, Patil K, Pfenniger A, Knight BP, Verma N. Implementation of an intravenous sotalol initiation protocol: Implications for feasibility, safety, and length of stay. J Cardiovasc Electrophysiol. 2023;34(3):502–506. doi: 10.1111/jce.15819.

[10] Steinberg BA, Holubkov R, Deering T, Groh CA, Mittal S, Kennedy R, Pokharel P, Perez M, Savona S, Verma N, Watt K, Piccini JP, Bunch TJ. Expedited loading with intravenous sotalol is safe and feasible-primary results of the Prospective Evaluation Analysis and Kinetics of IV Sotalol (PEAKS) Registry. Heart Rhythm. 2024;21(7):1134–1142. doi: 10.1016/j.hrthm.2024.02.046.

[11] Lakkireddy D, Ahmed A, Atkins D, Bawa D, Garg J, Bush J, Charate R, Bommana S, Pothineni NVK, Kabra R, Darden D, Koreber S, Tummala R, Vasamreddy C, Park P, Mohanty S, Gopinathannair R, Seo BW, Natale A, Kennedy R. Feasibility and Safety of Intravenous Sotalol Loading in Adult Patients With Atrial Fibrillation (DASH-AF). JACC Clin Electrophysiol. 2023;9(4):555–564. doi: 10.1016/j.jacep.2022.11.026

[12] Dong M, Fukuda T, Selim S, Smith MA, Rabinovich-Guilatt L, Cassella JV, Vinks AA. Clinical trial simulations and pharmacometric analysis in pediatrics: application to inhaled loxapine in children and adolescents. Clin Pharmacokinet. 2017;56(10):1207– 1217. doi: 10.1007/s40262-017-0512-x

